# Synaptic gene expression changes in frontotemporal dementia due to the *MAPT* 10+16 mutation

**DOI:** 10.1101/2024.04.09.24305501

**Authors:** Owen Dando, Robert McGeachan, Jamie McQueen, Paul Baxter, Nathan Rockley, Hannah McAlister, Adharsh Prasad, Xin He, Declan King, Jamie Rose, Phillip B Jones, Jane Tulloch, Siddharthan Chandran, Colin Smith, Giles Hardingham, Tara L Spires-Jones

**Author notes:** Co-corresponding authors: Contact:Prof Tara Spires-Jones OR Dr Owen Dando. These authors contributed equally (Owen Dando, Robert McGeachan).

## Abstract

Mutations in the *MAPT* gene encoding tau protein can cause autosomal dominant neurodegenerative tauopathies including frontotemporal dementia (often with Parkinsonism). In Alzheimer’s disease, the most common tauopathy, synapse loss is the strongest pathological correlate of cognitive decline. Recently, PET imaging with synaptic tracers revealed clinically relevant loss of synapses in primary tauopathies; however, the molecular mechanisms leading to synapse degeneration in primary tauopathies remain largely unknown. In this study, we examined post-mortem brain tissue from people who died with frontotemporal dementia with tau pathology (FTDtau) caused by the *MAPT* intronic exon 10+16 mutation, which increases splice variants containing exon 10 resulting in higher levels of tau with four microtubule binding domains. We used RNA sequencing and histopathology to examine temporal cortex and visual cortex, to look for molecular phenotypes compared to age, sex, and RNA integrity matched participants who died without neurological disease (n=12 per group). Bulk tissue RNA sequencing reveals substantial downregulation of gene expression associated with synaptic function. Upregulated biological pathways in human *MAPT* 10+16 brain included those involved in transcriptional regulation, DNA damage response, and neuroinflammation. Histopathology confirmed increased pathological tau accumulation in FTDtau cortex as well as a loss of presynaptic protein staining, and region-specific increased colocalization of phospho-tau with synapses in temporal cortex. Our data indicate that synaptic pathology likely contributes to pathogenesis in FTDtau caused by the *MAPT* 10+16 mutation.

## Introduction

Tau protein, encoded by the *MAPT* gene, is a microtubule binding protein that plays a role in many neuronal processes[23]. During the progression of diseases called tauopathies, tau protein becomes excessively phosphorylated and aggregates in pathological lesions in neurons, astrocytes, and oligodendrocytes. In addition to tau pathology, all tauopathies share the phenotype of progressive neurodegeneration. Over 60 mutations in the *MAPT* gene that cause neurodegenerative tauopathies have been identified [13], supporting a causative role for pathological changes in tau in causing degeneration.

In Alzheimer’s disease, the most common tauopathy, extensive evidence links synapse loss with cognitive decline and pathological tau has been observed within synapses in post-mortem brain samples [6, 15, 46]. However, Alzheimer’s disease is a so-called “secondary” tauopathy because changes in amyloid beta are thought to be the primary cause of disease, and amyloid beta oligomers are well known to cause synapse dysfunction and loss. Recent evidence from two “primary” tauopathies, progressive supranuclear palsy and corticobasal degeneration, using PET imaging with a presynaptic protein radioligand that binds SV2A indicate that synapse loss occurs in these diseases in the absence of amyloid pathology and correlates with symptom progression [17, 18]. Similarly, PET imaging of people with behavioural variant frontotemporal dementia (FTD) revealed synapse loss compared to controls which correlated with cognitive impairments [35]; however only half of the people with FTD have tau pathology in the brain (with TDP-43 or FUS accumulating in the other half)[37]. Relatively little is known about mechanisms of synapse degeneration in human FTD with tau pathology (FTDtau).

In order to better understand the mechanisms of neurodegeneration induced by tau, we examined post-mortem brain tissue from people with FTDtau caused by the *MAPT* intronic 10+16 mutation. This mutation increases splice variants containing exon 10 resulting in higher levels of tau with four microtubule binding repeats (4R tau) [12]. This mutation was first described in 1998 in people with frontotemporal dementia with Parkinsonism linked to chromosome 17,[19] and has subsequently been found in people with progressive subcortical gliosis,[14] behavioural variant FTD,[41] and a dementia symptoms clinically identical to Alzheimer’s type dementia.[9] The *MAPT* 10+16 mutation is associated with substantial neurodegeneration and accumulation of tau pathology in neurons and in some cases in astrocytes and oligodendrocytes.[9, 14, 41] In model systems, *MAPT* 10+16 expression is associated with synaptic dysregulation and degeneration. Genetically engineered mice with expressing human *MAPT* with the 10+16 mutation have age-dependent synapse loss and impaired synaptic plasticity,[47] and induced pluripotent stem cell derived neurons with the *MAPT* 10+16 mutation have pathological excitability.[22, 27]

Here we tested the hypothesis that the *MAPT* 10+16 mutation causes synaptic degeneration in people with FTDtau.

## Methods

### Human subjects

Use of human brain tissue was reviewed and approved by the contributing brain banks and the Academic and Clinical Central Office for Research and Development (ACCORD) and the medical research ethics committee AMREC a joint office of the University of Edinburgh and National Health Service Lothian, approval number 15-HV-016). *Post-mortem* brain samples were taken from Brodmann Area 41/42 (BA41/42) in the superior temporal gyrus (STG) and Brodmann Area 17 (BA17) in the primary visual cortex (VIS). These regions were chosen as the STG is affected early in the disease and has substantial pathology by end-stage, whereas the visual cortex exhibits minimal pathology even in the late stages of disease. Inclusion criteria for FTDtau cases were genetic conformation of the *MAPT* 10+16 mutation (undertaken by brain banks from which tissue was acquired), a clinical diagnosis of dementia, and a neuropathological diagnosis of FTD with tau pathology. Control cases were age and sex matched and included based on having no clinical diagnoses of neurological or psychiatric diseases. The exclusion criteria for both groups were accumulation of amyloid beta, alpha-synuclein, TDP-43, or tau (in controls only) in levels beyond that expected for normal ageing. All cases were blinded throughout experimentation and data acquisition, until the statistical analysis of data. Summary demographics of brain tissue donors are included in **Table 1**.

**Table 1:** Demographics of subjects. NS = not significant, * p < 0.05.

| RNAseq Study (Figures 1&2) | Control (N=12) | FTDtau (N=12) | Difference between groups |
| --- | --- | --- | --- |
| **Age** - Mean (SD) | 62.3 (9.27) | 60.9 (6.32) | NS (t = 0.14, df = 17.83, p = 0.89) |
| **Sex** (F) | 5 (41.7%) | 5 (41.7%) | NS ($\chi^2$ squared=0, df=1, p=1) |
| **RIN** - Mean (SD) | 6.28 (0.79) | 5.95 (0.98) | NS (t = 1.69, df = 20.23, p = 0.11) |
| IHC study (Figure 3) | Control (N=11) | FTDtau (N=11) |  |
| **Age** - Mean (SD) | 63.6 (9.97) | 61.4 (7.03) | NS (t = 0.62, df = 17.97, p = 0.54) |
| **Sex** (F) | 5 (45.5%) | 5 (45.5%) | NS ($\chi^2$ squared=0, df=1, p=1) |
| IHC study (Figures 4&5) | Control (N=9) | FTDtau (N=7) |  |
| **Age** - Mean (SD) | 63.7 (9.23) | 62.6 (7.68) | NS (t = 0.26, df = 13.9, p = 0.80) |
| **Sex** (F) | 3 (33.3%) | 3 (43.9%) | NS ($\chi^2$ squared=0, df=1, p=1) |
| RNAseq age study (Figure S1) | Young control (<65) (N=10) | Older control (>65) (N=11) |  |
| **Age** - Mean (SD) | 46.9 (8.96) | 79.6 (10.7) | *(t = -7.62, df = 18.89, p < 0.0001) |
| **Sex** (F) | 3 (30%) | 6 (54.5%) | NS ($\chi^2$ squared=0.48, df=1, p=0.49) |
| **RIN** - Mean (SD) | 6.58 (0.63) | 6.44 (0.99) | NS (t = 0.40, df = 17.10, p=0.69) |

### RNA sequencing

Total RNA was extracted from frozen human brain samples using the RNeasy Lipid Tissue kit (Qiagen) as per the manufacturer’s instructions, including spin-column DNAse-treatment. RNA integrity number (RIN) was measured on an Agilent 2100 Bioanalyzer using the Agilent RNA 6000 Nano Kit, with the median RIN values: 6.25 (controls) and 6.05 (FTD-Tau) (Table 1). RNA-seq libraries were prepared for next-generation sequencing by Edinburgh Genomics using TrueSeq Stranded Total RNA V2 library preparation. The libraries were pooled and sequenced to 75 base paired-end, to a depth of ∼35 million paired-end reads per sample on the Illumina HiSeq 4000 platform. RNA-seq reads were mapped to genome sequences using STAR (Spliced Transcripts Alignment to a Reference [8]). Per-gene read counts were summarised using featureCounts,[32] and differential expression analysis performed using DESeq2,[34] with a significance threshold at a Benjamini-Hochberg-adjusted *P* value of <0.05. Gene ontology analyses were performed with topGO.[1]

### Immunohistochemistry staining and imaging

Tissue sections were dewaxed with Xylene and rehydrated with decreasing concentrations of Ethanol-to-water solutions. For examination of tau burden, tau was stained with AT8 antibody (recognising tau phosphorylated at ser202/thr205, ThermoFisher MN1020, AB_223647) and visualised with the Leica Novolink Polymer Detection Kit. Tissue was counterstained with haemotoxylin to label nuclei and coverslips were mounted using DPX. Stereology was carried out on the stained slides using a Zeiss Axioimager Z2 and StereoInvestigator software (MicroBrightField). Neurofibrillary tangle counts were carried out using the Optical Fractionator workflow in StereoInvestigator and AT8 burden was calculated by automated detection of AT8 staining within grey matter.

For immunofluorescences staining of synapses, tau, and astrocytes, after dewaxing and rehydration, tissue was subjected to citrate buffer (pH6.0) antigen retrieval in a pressure cooker for 3 minutes. Sudan Black was used to limit autofluorescence. Tissue sections were permeabilized (1xPBS, 0.3% Triton X-100), blocked (1xPBS,0.3% Triton-X-100, 10% normal donkey serum), and incubated overnight in primary antibodies to GFAP (Abcam ab4674, 1:5000) and AT8 (Thermo Fisher MN1020, 1:500). The next day sections were washed and incubated in secondary antibodies (goat anti-chicken IgGY AlexaFluor 405, Thermo Fisher A48260, 1:500 and goat anti-mouse IgG1 AlexaFluor647, Thermo Fisher A21240, 1:500) followed by rinsing and incubation in direct labelled anti synaptophysin antibody (AlexaFluor488, Abcam ab8049, 1:250). Images were acquired using a confocal microscope (Leica TCS8) with a 63x oil immersion objective. Ten image stacks were taken for each case and brain region sampled randomly through all 6 cortical layers. Laser and detector settings were kept constant for each individual sample but were adjusted accordingly on a case-by-case basis to account for variability in stain quality. Using a custom MATLAB script, the image stacks were segmented, and a custom python script was used to calculate the volume of colocalization between channels in the segmented images. We observed that a subset of cases exhibited notable variability in tissue preservation and immunostaining quality. To ensure our data was accurate and representative, we implemented exclusion criteria for cases showing signs of tissue architecture degradation and/or inadequate immunostaining

### Statistical analyses

Statistical analyses were performed in R version 4.3.2 with R studio. Demographic characteristics were analyzed with t-test, Wilcoxon test, or Chi-squared test as appropriate for the data type and distribution (assessed with Shapiro-Wilks tests and inspection of histograms). Tau burdens measured with stereology were assessed using general linear models to evaluate the effect of diagnosis, brain region, and sex. Statistical analysis of immunofluorescence data was performed using linear mixed effects models with diagnosis, brain region and sex as fixed effects and case as a random effect to avoid pseudoreplication. To test if the data met the assumptions of equal variance and normality, residual plots were visually examined. Additionally, Shapiro-Wilks tests were used to corroborate the normality of the data. If the data did not meet the assumptions, Tukey transformations were performed to better fit the assumptions of normality and equal variance. ANOVAs were performed on the linear mixed effect models using Satterthwaite’s method. Tukey corrected post-hoc comparisons of means were performed on factors that were found to significantly affect the data. All contrast estimates, F statistics, t statistics and p values are reported as Tukey transformed values where transformed. Boxplots and graphs were generated in RStudio. Boxplots show untransformed data to demonstrate variability between in the raw data, with each data point representing the median or the mean for an individual human case. The distribution of the data was plotted to determine if the median or mean was chosen.

### Data availability statement

Human post-mortem RNAseq results will be deposited in an appropriate online repository before publication. Spreadsheets of IHC data and all statistical analysis scripts can be found on the University of Edinburgh DataShare repository https://datashare.ed.ac.uk/handle/10283/3076 (DOI to be added upon acceptance for publication). IHC image analysis software is available online here https://github.com/Spires-Jones-Lab/3DBurdenn_Python, and raw microscope images are available from the corresponding author upon reasonable request.

## Results

### Gene expression changes in FTDtau *MAPT* 10+16 brain tissue

Bulk RNA sequencing was used to examine gene expression changes in brain tissue from people with FTDtau due to the *MAPT* 10+16 mutation controls. Our cases and controls (n=12 per group) were matched for age, sex, and RNA integrity (**Table 1**). Examination of the RNAseq data from superior temporal gyrus and visual cortex (STG and VIS) shows substantial gene expression changes in FTDtau cases with adjusted p value < 0.05 (**Figure 1A, B**). Using the synaptic gene ontologies database (SynGO[26]), we observe transcripts associated with pre and post-synaptic localisation and function are changed in FTDtau (**Figure 1B, C**). When filtered to look at differentially expressed genes altered by more than 1 fold (doubled or halved in FTDtau) with an adjusted p value <0.05, we observe 128 upregulated and 63 downregulated transcripts in STG and 686 upregulated and 614 downregulated transcripts in VIS.

**Figure 1:**
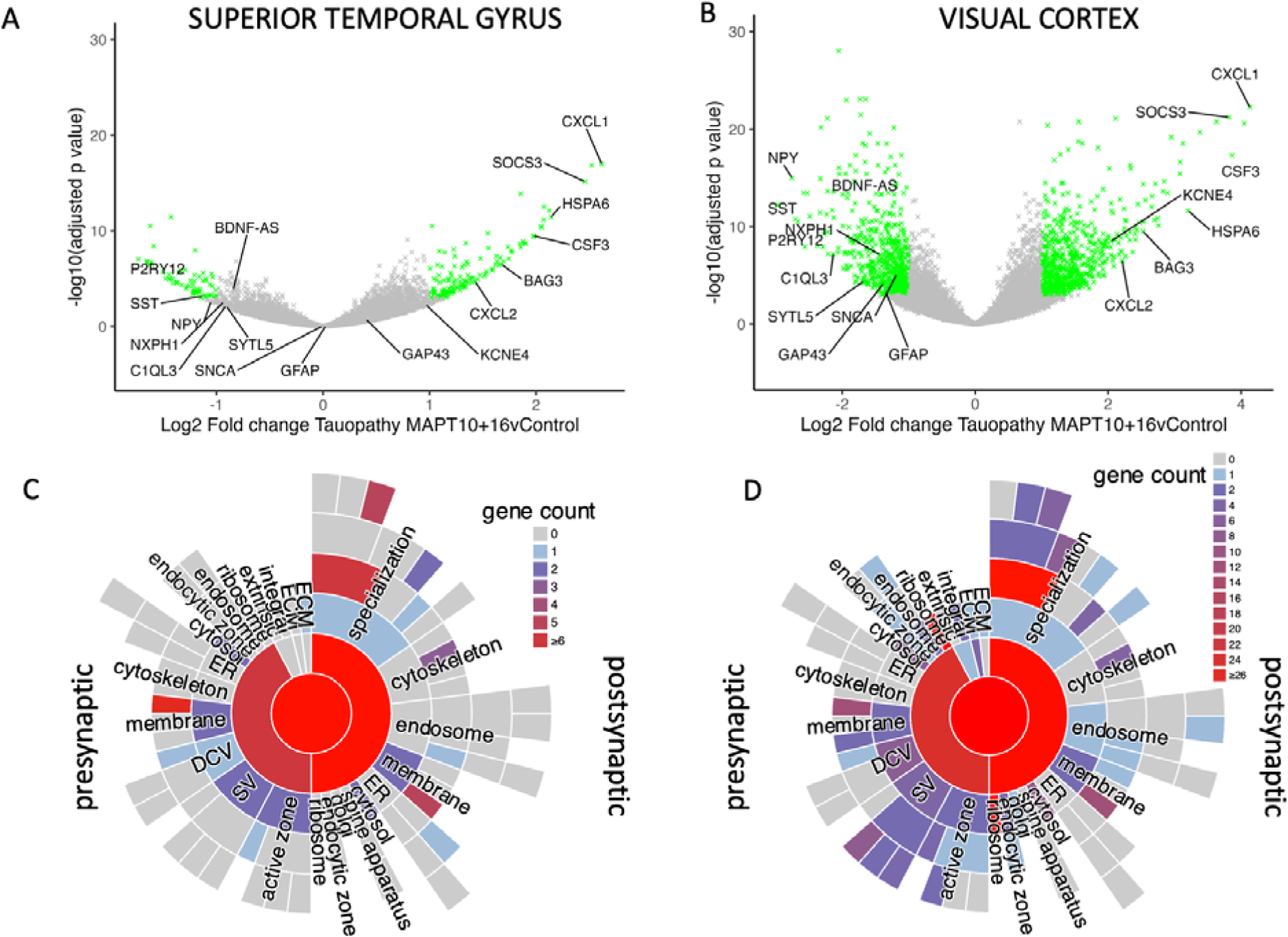
Changes in gene expression in FTDtau. Volcano plots of RNAseq data show changes in gene expression in FTDtau in both superior temporal gyrus (A) and primary visual cortex (B). Each cross represents an individual transcript. Genes with a log2 fold change >1 or <-1 and adjusted p value <0.05 are coloured green and transcripts of interest are labelled. SYNGO analysis of all significantly changed genes (p adj <0.05) shows that in both STG (C) and VIS (D) substantial numbers of both pre- and post-synaptic transcripts are changed in FTDtau compared to control cases.

Gene Ontology analysis of biological processes changed in FTDtau vs controls reveals downregulation of many pathways involved in synaptic function including synaptic transmission, synaptic plasticity, regulation of NMDA receptor activity, synapse assembly and learning (**Figure 2**). GO analysis from organoids with the *MAPT* 10+16 mutation compared to isogenic controls show downregulation of 10 pathways identical to those observed in post-mortem tissue (**Figure 2**). In human post-mortem FTDtau tissue, upregulated transcripts included several pathways involved in regulation of transcription and cellular damage response or cell cycle regulation. There were regional differences in pathways involved in inflammation with many more inflammatory pathways upregulated in VIS than STG (**Figure 2**). When we expanded the number of controls beyond our age-matching range for FTDtau cases and compared old (>65 years old, n=11) vs young (<65 years old, n=10) controls, we observe similar decreases in synaptic gene expression and increases in pathways involved in inflammation, transcription, and cell cycle control (**Supplementary Figure 1**). Interestingly, both the inflammatory upregulation and synaptic downregulation are more pronounced in STG than VIS in old vs young controls in contrast to higher inflammation in VIS when comparing FTDtau cases to controls. Together, these RNAseq data indicate substantial downregulation of synaptic genes and upregulation of transcriptional regulation and damage response.

**Figure 2:**
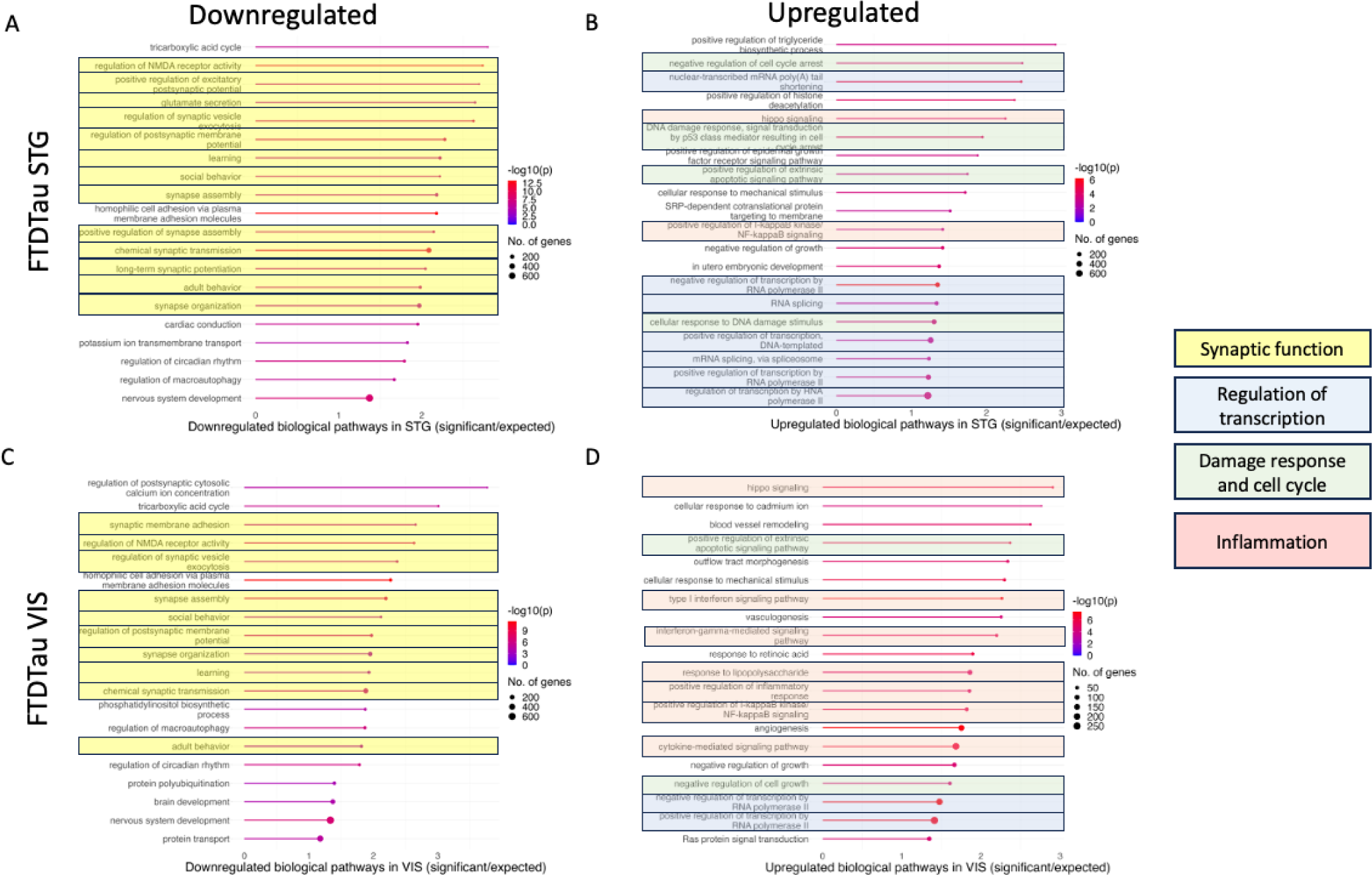
The top 20 down-regulated (A, C) and up-regulated (B,D) pathways in Gene Ontology analysis of RNAseq data reveal that in both STG (A, B), and visual cortex (C, D), expression of genes involved in synaptic function are decreased (highlighted in yellow) and genes involved in regulation of transcription (blue), damage response or cell cycle regulation (green), and inflammation (pink) are increased in FTD-tau 10+16 compared to controls.

### Immunohistochemistry confirms loss of synaptic protein and synaptic accumulation of pathological tau in FTDtau brain

To further explore pathological changes in FTDtau *MAPT* 10+16 carriers, we performed immunohistochemistry on formalin fixed, paraffin-embedded brain tissue. Staining for pathological tau phosphorylated at ser 202 and thr 205 using AT8 antibody confirms tau pathology in FTDtau both when examining the burden of AT8 staining (percent tissue area occupied by staining) and when counting neurofibrillary tangles (NFTs). We observe the expected increase in tau pathology in FTDtau compared to control and the expected higher levels of pathology in STG compared to VIS (**Figure 3**).

**Figure 3:**
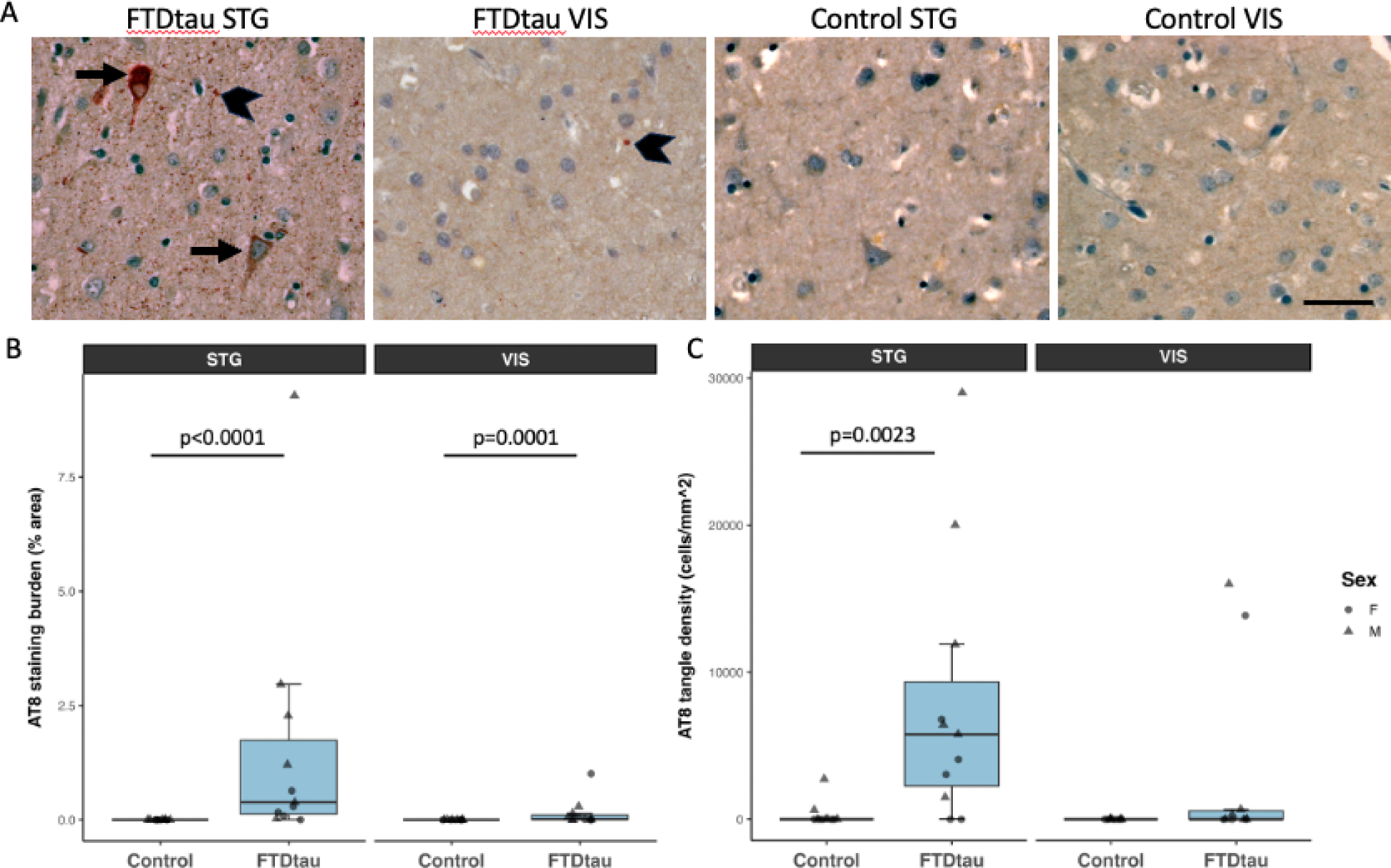
Tau pathology in FTD-tau. **A.** Immunohistochemistry staining of phospho-tau with AT8 antibody (red/brown) and nuclei (hematoxilyn – blue counterstain) shows AT8 accumulation in tangles in neuronal cell bodies (arrows) and in the neuropil (chevrons). **B.** AT8 staining burde (% of cortex occupied by AT8 staining) is increased in FTD-tau compared to control cases (ANOVA after linear mixed effects model F[1,20]=47.06, p<0.0001) and increased in superior temporal gyrus compared to visual cortex (F[1,20]=23.17, p=0.0001). **C.** Density of AT8 positive tangles is similarly increased in FTD-tau (F[1,20]=7.16, p=0.014) and higher in STG than VIS (F[1,20]= 4.62, p=0.044). n=11 FTD-tau and 11 control in this experiment. *p* values on graphs are from post-hoc Tukey-corrected pairwise tests. Scale bar represents 20 μm.

Immunofluorescence staining of AT8, synaptophysin, P2Y12, and GFAP was used to investigate presynaptic proteins and the colocalization of synapses with astrocytes, microglia, and tau pathology (**Figure 4**). The percent volume of tissue occupied by synaptophysin staining in confocal image stacks was significantly reduced in FTDtau compared to controls (**Figure 5**), supporting the observation in RNAseq data of decreases in transcripts important for synaptic structure and function. AT8 burden in the confocal image stacks was increased in FTDtau compared to control as expected. We observed colocalization of phospho-tau with astrocytes as has been previously published in FTD but surprisingly, we did not observe any increased astrogliosis or any change in colocalisation of synaptic protein with astrocytes in FTDtau compared to control brain tissue. In line with GFAP staining in our IHC Study, we do not see changes in GFAP in STG in our RNAseq data (log2 fold change 0.02, p=0.98), while in visual cortex, GFAP levels were significantly decreased (log2 fold change −1.35, p=0.04). Similarly, we observed reduced staining of microglia with P2Y12 (Figure 5), which was supported by the bulk change −2.67, p<0.0001). We observed AT8 staining within microglia increased in FTDtau compared to control but decreases in colocalization of synaptophysin in P2Y12 positive microglia.

**Figure 4:**
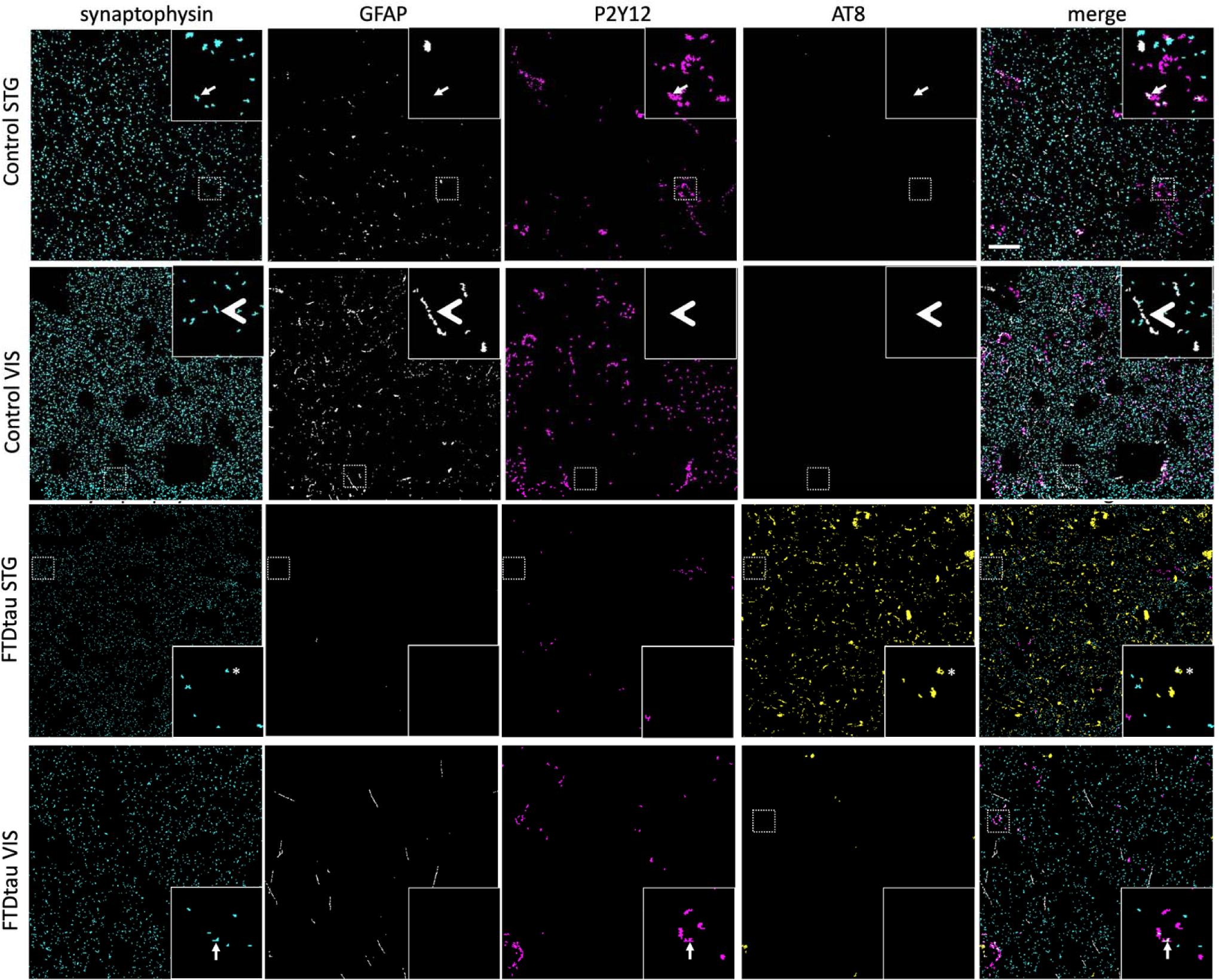
Immunofluorescence staining and confocal imaging were used to examine colocaliztion between synaptophysin (cyan), GFAP (grey), P2Y12 (magenta) and AT8 (yellow). Large panels are maximum intensity projections of 10 sections of segmented confocal stacks. Insets are 10 μm x 10 μm regions of interest from a single confocal section. Arrows show synaptophysin and P2Y12 colocalization, arrowheads show synaptophysin and GFAP colocalization, asterisk shows synaptophysin AT8 colocalization. Scale bar represents 20 μm.

**Figure 5:**
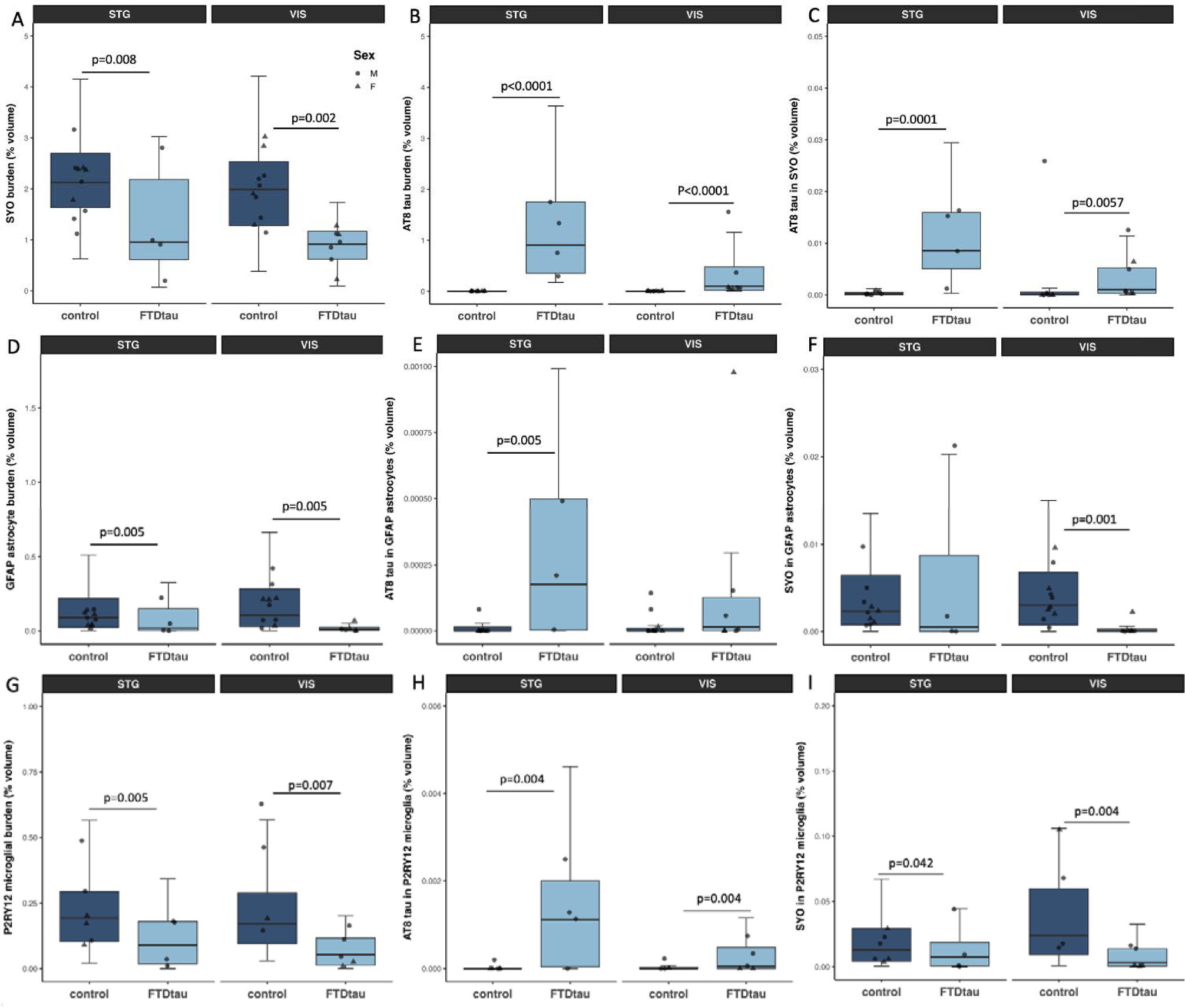
Analysis of confocal images shows a reduced volume of the cortex occupied by synaptophysin staining in FTDtau compared to controls in both superior temporal gyrus and visual cortex (A, ANOVA effect of diagnosis F[1,15.61]=12.02, p=0.003). Cortical volume occupied by AT8 was increased in both brain regions in the confocal stacks from FTDtau cases as expected (B, ANOVA effect of diagnosis F[1,16.39]=98.10, p<0.0001). Colocalisation between AT8 and synapophysin was increased in FTDtau (C, ANOVA effect of diagnosis F[1,13.77]=19.87, p=0.0006). We observed decreases in reactive astrocyte staining with GFAP in FTDtau (D, ANOVA effect of diagnosis F[1,14.38]=20.03, p=0.0005), and an increase in AT8 tau colocalised in astrocytes in FTDtau (E, ANOVA effect of diagnosis F[1,14.61]=9.05, p=0.009). There was a decrease in colocalization of synaptophysin and GFAP in FTDtau (F, ANOVA effect of diagnosis F[1,14.70]=6.67, p=0.02). Microglial burdens measured by P2RY12 staining are decreased (G, ANOVA effect of diagnosis F[1,13.21]=11.07, p=0.005), colocalization of AT8 tau within microglia is increased (H, ANOVA effect of diagnosis F[1,13.00]=8.80, p=0.011), and the colocalization of synapses in microglia is decreased in FTDtau (I, ANOVA effect of diagnosis F[1,13.98]= .27, p=0.012). *p* values on graphs are from Tukey corrected post-hoc tests after linear mixed effects models.

## Discussion

Our data are consistent with the hypothesis that the *MAPT* 10+16 mutation causes synaptic degeneration. We observe decreased synaptic gene expression both in post-mortem brain tissue and in organoids expressing the *MAPT* 10+16 mutation. Recent data from *MAPT* 10+16 organoids similarly demonstrates substantial dysregulation of synaptic gene expression in neurons,[2] indicating that the decreases in synaptic gene expression we observe are not solely due to loss of neurons at end stages of disease. Pathways and transcripts involved in both excitatory and inhibitory signalling are altered in our dataset. In FTDtau *MAPT* 10+16 brain samples, we see downregulation of several pathways important for excitatory synapses including regulation of NMDA receptor activity, positive regulation of excitatory post-synaptic potential and long-term synaptic potentiation. There is a large body of evidence linking tau pathology to excitatory neurons. In AD, tau pathology accumulates early in excitatory neurons of the entorhinal cortex and hippocampus,[4, 20] and single nucleus sequencing data from post-mortem AD brain indicates tangles preferentially form in excitatory over inhibitory neurons.[40] Similarly, tau pathology accumulates in some excitatory pyramidal neurons in primary tauopathies including Progressive Supranuclear Palsy (PSP); however other neuronal subtypes (dopaminergic etc) and glia also accumulate substantial tau pathology in PSP.[29] We have observed excitatory synapse loss and tau accumulation within excitatory synapses in AD and PSP brain.[6, 6, 24, 25, 38]

In mouse models of tauopathy, tau and neurodegeneration affect predominantly in excitatory neurons.[10, 11] Excitatory synapses are also lost in these models,[28, 42] and functional changes are observed in excitatory neurons including longer down states and reduced firing rates[39] and hypoactivity of neural cicruits.[5] In human iPSC derived organoids expressing *MAPT* with the V337M mutation which causes FTD, excitatory neurons are lost over time and *ELAVL4*, a major regulator of glutamatergic cell fate involved in synaptic plasticity, was increased in V337M organoids compared to isogenic controls.[3] Interestingly, in our RNAseq data, *ELAVL4* was significantly decreased in both brain regions examined.

We also observed decreases in transcripts involved in inhibitory synapse function in our human post-mortem brain RNAseq study. SST, GABARB2, and GAD2 were all significantly decreased in FTDtau brain indicating a loss of inhibitory synapse function. Although not previously reported for FTDtau cases, this loss of inhibitory function is in line with previous studies of AD brain and mouse models of tauopathy. We have previously observed loss of inhibitory neurons and inhibitory synapse loss around plaques in human AD brain.[30] rTg4510 mice which express P301L mutant tau associated with FTD also have early loss of inhibitory synapses [44].

PCDH5, PCDHAC2, and PCDHGC4 are also among the top 5 downregulated transcripts in FTDtau brain in our study. These all encode protocadherin proteins important in neuronal cell adhesion and development. *PCDHGC4* variants have been linked to a neurodevelopmental disorder characterized by microcephaly, seizures, and intellectual disability [21]. The seizure phenotype may be due to an important role for protocadherins in apoptosis of cortical interneurons during development.[36] While most data on protocadherins and synapses relates to neurodevelopment, our data indicate they may also be involved in synapse degeneration.

Based on the accumulation of tau pathology in astrocytes and the essential roles astrocytes play in synaptic function, we hypothesized that astrocyte changes might contribute to synapse loss in FTDtau. In Alzheimer’s disease brain, we have previously observed substantial neuroinflammation in the synaptic proteome[16] and engulfment of synaptic proteins by astrocytes and microglia around plaques.[45] Amyloid pathology is well known to drive neuroinflammation in models of AD; however inflammatory complement activation is also observed in primary tauopathy brain and mouse models of tauopathy [7, 33]. Here we did not observe increased expression of complement related pathways in FTDtau; however we did observe increases in hippo signalling which plays a role in controlling inflammation [48]. We also see increases in regulation of NF-kappaB signalling, interferon signalling, and cytokine mediated signalling, all implicating neuroinflammation in FTDtau as has been previously suggested in tauopathies [31]. Interestingly, the inflammatory changes were more pronounced in visual cortex in our post-mortem samples indicating this could be an early response to tau accumulation as this brain region is affected late in disease and has less tau accumulation than temporal cortex

iPSC astrocytes expressing *MAPT* 10+16 have been shown to have elevated 4R tau levels, which likely affect their function.[43] Our previous data also suggest that GFAP positive astrocytes engulf synapses in both AD and PSP, which likely contributes to observed synapse loss.[6, 38] In AD but not PSP, we observe increases in synaptic protein inside P2Y12 positive microglia.[6, 38] Here we surprisingly observed decreased colocalization between synaptic proteins and both GFAP positive astrocytes and P2Y12 microglia in FTDtau, indicating potentially diverging roles in synapse-glia interaction between neurodegenerative tauopathies. However, it is worth noting the limited sample size used in these analyses due to the scarcity of available issue.

Together, our data indicate that synapse loss is an important phenotype in in frontotemporal dementia with tau pathology caused by the *MAPT* 10+16 mutation. Further investigation to prevent pathological tau induced synaptic dysfunction and loss will be important to develop therapeutic strategies.

## Acknowledgements

We gratefully acknowledge the brain tissue and iPSC donors and their families, without whom this work would not be possible. We also acknowledge Edinburgh Neuroscience and the FENS-Kavli Network of Excellence for facilitating collaborations leading to this work, the Alzheimer’s Scotland Dementia Research Centre for coordinating post-mortem brain tissue donations, and Dr Kathryn Bowles for useful discussions of the work. Human tissue samples were provided by the Edinburgh Brain Bank, Manchester Brain Bank (which is part of the Brains for Dementia Research Initiative, jointly funded by Alzheimer’s Society and Alzheimer’s Research UK), MRC London Neurodegenerative Diseases Brain Bank, and the Neurodegenerative Disease Brain Bank at the University of California, San Francisco, which receives funding support from NIH grants P01AG019724 and P50AG023501, the Consortium for Frontotemporal Dementia Research, and the Tau Consortium. This work was supported by the UK Dementia Research Institute [award number UK DRI-Edin005] through UK DRI Ltd, principally funded by the UK Medical Research Council. The confocal microscope was generously funded by Alzheimer’s Research UK (ARUK-EG2016A-6) and a Wellcome Trust Institutional Strategic Support Fund at the University of Edinburgh.

## Competing Interests

TSJ is on the Scientific Advisory Board of Race Against Dementia, Alzheimer’s Research UK, and Cognition Therapeutics, and has consulted for Jay Therapeutics. None of these had any influence over the current paper.

---

**Supplementary Figure 1:**
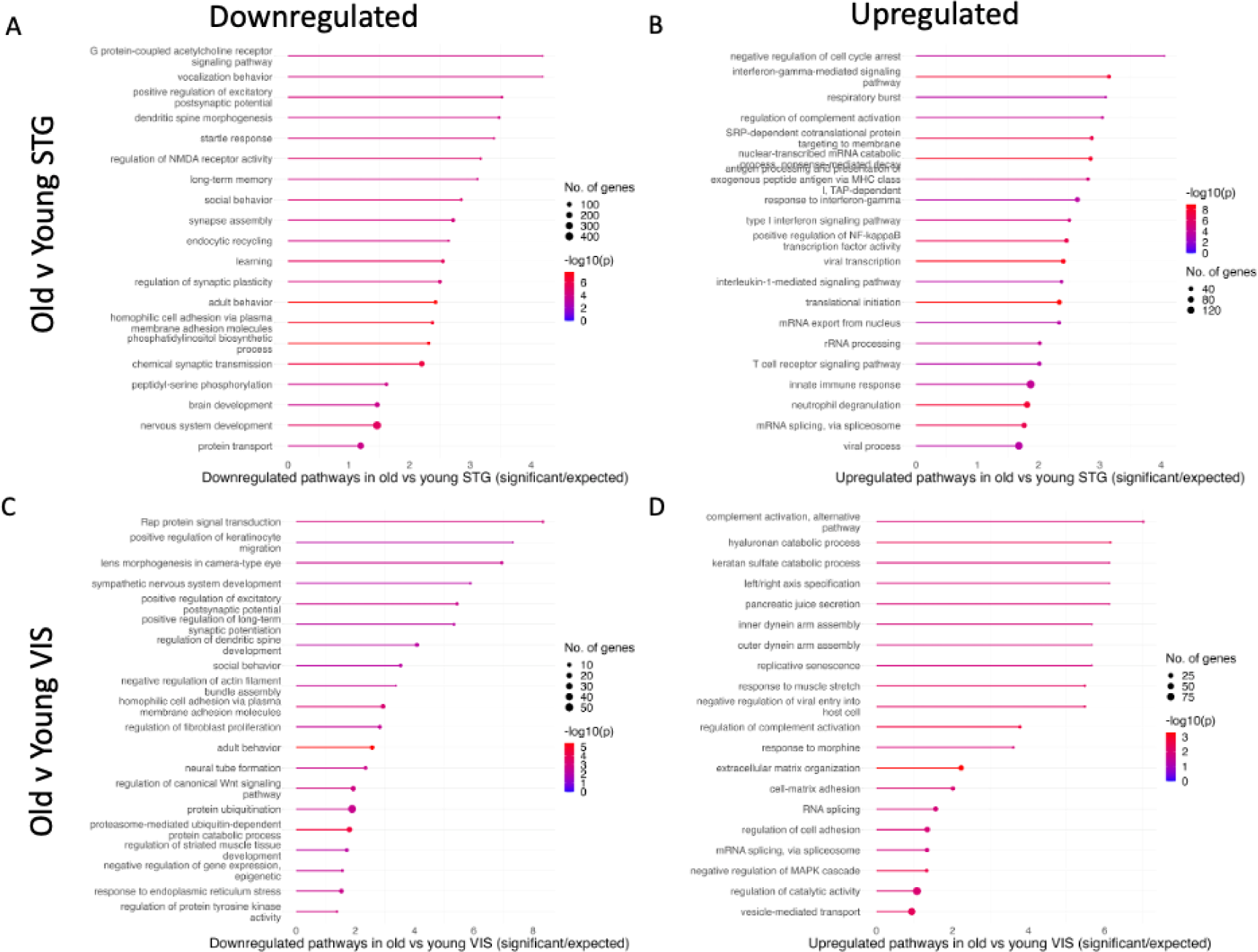
When comparing older vs younger controls (cutoff 65 years of age), the top 20 down-regulated (A, C) and up-regulated (B,D) pathways in Gene Ontology analysis of RNAseq data reveal that in both STG (A, B), and visual cortex (C, D), expression of genes involved in synaptic function are decreased and genes involved in inflammation and regulation of the cell cycle are increased.

## Notes

### Author Declarations

Use of human brain tissue was reviewed and approved by the contributing brain banks and the Academic and Clinical Central Office for Research and Development (ACCORD) and the medical research ethics committee AMREC a joint office of the University of Edinburgh and National Health Service Lothian, approval number 15-HV-016).

